# CO-INFECTS: A Highly Affordable, Portable, Nucleic-Acid-Based Rapid Detector of Active Respiratory Co-Infections via “Swab-to-Result” Integration

**DOI:** 10.1101/2024.08.10.24311776

**Authors:** Sudip Nag, Saptarshi Banerjee, Aditya Bandopadhyay, Indranath Banerjee, Subhasis Jana, Arindam Mondal, Suman Chakraborty

## Abstract

Early diagnosis of active viral co-infections of similar symptomatic pattern is critical for preventing life-threatening respiratory complications. While facilities for the same are currently available, those are either over-expensive or not amenable for deployment in resource-limited settings. We report a first of its kind isothermal colorimetric nucleic acid-based test integrated in a user-friendly, portable device, that can differentially diagnose respiratory viral infections having similar symptomatic pattern, and illustrate its performance by detecting co-infections of SARS-CoV-2 with seasonal influenza A virus from unprocessed swab sample in an extraction-free analysis mode within 45 minutes without intermediate manual steps. The test obviates the need of any complex disposable cartridge in preference to common laboratory utilities. Further, it harnesses machine learning empowered and smartphone interfaced colorimetric analysis without needing any specialized optical detector. The efficacy of the test is validated via community-adapted randomized field trials at underserved rural settings by frontline health workers, reporting sensitivity >96% and specificity > 98% in 1500 cohorts. The method in the form of three test genes with one internal control assay evidences to facilitate viral surveillance and enables diagnosing complex respiratory infections early enough in a plethora of settings ranging from general hospitals to the community centres, overcoming barriers to access of high-end facilities. Such adaptation of scientifically supreme multiplex nucleic acid based diagnostics in the framework of a simple and user friendly rapid test appears to be the future of infectious disease management in a democratized framework, eradicating the disparities in affordability and accessibility in resource-limited settings.

## 1. Introduction

The onset of novel coronavirus pandemic, despite its devastating consequence, epitomized a landmark in setting up new vistas of infectious disease detection and management. Thanks to the highly aggressive research and development activities for overcoming the crisis, the pandemic challenge fostered new opportunities of developing the foundation of several interventions and measures having implications significantly more far-reaching than the immediate interest of pandemic control itself. Post the peak phases of the pandemic, strict measures of personal hygiene (use of face mask and sanitization) and physical distancing cum isolation resulted in a temporary decline in the deadly impacts of different other respiratory viruses as well (1–7). However, as these measures continued to be relaxed, threatening co-infections of multiple viruses silently re-emerged to become a new threat, with COVID-19 being added to the spectra of the previously known invading pathogens. The challenge compounded many folds for co-morbid or immune-compromised patients or the ones having previous episodes of serious respiratory ailments. Despite the emergence of extremely inventive diagnostic tests in response to the recent pandemic crisis, there seems to be no practical answer to this newly emerged challenge except for a few privileged ones who can afford to have timely access to high-end medical facilities.

The lethal aspects of co-infections of SARS-CoV-2 with other endemic respiratory viruses came to the spotlight as hospitalization and critical care became the need of the hour during the pandemic peaks (8,9). Since influenza (common flu) is known to affect a large population seasonally, it logically appeared to be a major contributor to such co-infection challenge (10). Currently, influenza affects around a billion individuals annually, culminating into 3–5 million cases of severe illness and 290000 to 650000 respiratory deaths (https://www.who.int/news-room/fact-sheets/detail/influenza-(seasonal)), especially in immunocompromised people or those with comorbidities, or amongst individuals co-infected by other respiratory pathogens. While testing for influenza virtually does not exist in the public health arena (11), its consequences could have overlapping symptomatic manifestations akin to COVID-19 such as cough, cold, fever, head- and body-ache, sore-throat (12,13), similar mode of transmissibility via respiratory droplets and contaminated interfaces, analogous inflammatory responses that may compound to result in severe damage of the lung-epithelial cells (14–23), and potential threat to result in lethal secondary infections like severe pneumonia causing life threatening complications. Notably, while in isolation COVID-19 or influenza may induce only limited damage to most patients, their concomitant action may amplify the clinical complications largely. Such co-infected patients were thus reported to show reduced level of severe lymphopenia in peripheral blood resulting in hampered neutralizing antibody (Nab) and T cell responses (24), higher levels of cytokines and chemokines than a single infection condition and prolonged viral particle shedding. Clinically, this could be evidenced by enhanced disease severity and lung injury, with higher probabilities of requiring non-invasive or invasive respiratory support (ventilation) as a life-saving measure (25,26).

With such imperative implications of co-infection of different respiratory viruses, their testing appears to be central in isolating at-risk patients having challenged responses to immunomodulatory and antiviral therapy. In resourced-hospital and high-end diagnostic centres, respiratory samples collected for this purpose from symptomatic individuals may be subjected to multiplex reverse transcription-quantitative polymerase chain reaction (RT-qPCR) testing for a panel of respiratory viruses for which genomic sequence information of the pathogen is available. This commonly includes influenza A (H1-pdm09 and H3 subtypes) and B viruses, RSV (Respiratory syncytial virus)- A and B, human coronaviruses (hCoV-HKU1, 229E, OC43, NL63), human metapneumoviruses (hMPVs), human entero/rhinoviruses (hERVs), parainfluenza viruses (PIV 1-4) and adenovirus, for which the target genes for PCR-based tests are established. In the SARS-CoV-2 pandemic, the early availability of the genome (27) facilitated the development of reliable primer sets for developing its PCR based tests (28,29). However, while scientifically there is no restriction in using such PCR-based procedure for the differential diagnostics of respiratory viral infections, this alone remained inadequate for addressing the diagnostic challenge because the high-end facilities needed for such tests appeared to be scarce as compared to the exceeding demand (30).

The above limitation in RT-qPCR test capacity prompted for the development of independent and affordable alternative nucleic acid-based tests, even for detecting single infections. The reverse-transcription loop-mediated isothermal amplification (RT-LAMP) test thus emerged as an attractive substitute as it eliminates the needs for thermocyclers so that a vast range of isothermal heating instruments could be used instead (31), alongside unrestricted usability of the reagents having expired patents (32). Commonly, in the point of care (POC) LAMP-based tests, the conventional fluorescence-based detection systems were replaced with drastically simplified colorimetric detection modes with the aid of pH-sensitive dyes (33,34). The COVID-19 pandemic provided opportunities of evaluating such different RT-LAMP based assays for their efficacy (34–39). A few of these tests were developed in semi-quantitative multiplexed framework as well (40), presenting their viable propositions as significantly more accurate alternatives to the common rapid antigen tests (41).

However, despite all promises, the development of LAMP-based diagnostic tools could not match with the outstanding needs of infectious disease control. This could be attributed to their limitations in field adaptation in uncontrolled settings under the threat of carryover contamination, limited or no multiplexing, and complex primer design, among others. To satisfy the requirements of accuracy (high clinical sensitivity and specificity), simplified variants of the LAMP-based tests more commonly had to be discarded, whereas even a reasonable trade-off between simplicity and accuracy remained hard to achieve. Somewhat less sensitive yet affordable rapid antigen-based screening tests thus tended to fill up this void of the-then fertile COVID-19 diagnostic market (42). Evidently, more decisive POC tests having reasonably high accuracy and the capability of detecting early infections including co-infections remained to be an unmet need for eliminating any unwarranted clinical dilemma as well as hit-and-miss therapies (43). In an attempt to address the problem, an electrochemical genomagnetic assay has recently been established (44) to detect viral co-infection. However, this test could not meet the expectation in terms of sensitivity, specificity as well the turnaround time. Furthermore, the detection of co-infection was reported in very small cohorts of healthy and diseased subjects. Apparently inventive solutions such as folded paper-based kits (45) were indeed put forward to this end, although questionable robustness of fabrication and operation made them unreliable for use in professional diagnostic setups as against academic research labs. From practical considerations, since complete instrument-free nucleic acid based testing would lose many non-negotiable process controls, the most pragmatic wish list would still include minimal basic-equipment as hardware with fully integrated thermal, electronic and readout units for easy adopting across different public-health settings, with the value-added proposition of accommodating multiple tests for ruling out the possibilities of different infections that may otherwise express with similar symptoms.

Responding to the above need, POC variants of different LAMP-based tests for multiplex targets off late emerged with great promises (46–48) including the co-infection of SARS-CoV-2 with other respiratory bacterial infections (49). This could reflect in unprecedented market growth of 18.5% in multiplex respiratory tests from 2021 to 2022 in the USA alone, despite an unfavourable price escalation of 10.4% (https://www.medicaldevice-network.com/analyst-comment/poc-respiratory-multiplex-tests/?cf-view). Several popular product lines such as such Xpert® Xpress CoV-2/Flu/RSV plus (https://www.cepheid.com/en-GB/tests/respiratory/xpert-xpress-cov-2-flu-rsv-plus.html), The BioFire® Respiratory 2.1 (RP2.1) Panel (https://www.biofiredx.com/products/the-filmarray-panels/#respiratory), and the LuciraCOVID-19 & Flu Test (https://checkit.lucirahealth.ca/products/lucira-covid-19-flu-test; https://www.fda.gov/media/165690/download) thus entered into the POC diagnostic market akin to consumer electronic products. The latter one, which is a single use (disposable) test kit intended for simultaneous rapid in vitro qualitative detection and differentiation of SARS CoV-2, influenza A, and influenza B viral nucleic acid from anterior nasal swab samples, is of particular relevance to the present context, but was priced unfavourably (around 70 US$ per test). Summarily, while these commercial kits could signify landmarks in bringing high-end nucleic acid based tests for detecting respiratory co-infections towards off-the-shelf availability, challenges continued to persist either because of their prohibitively high cost or complexities, limitations in terms of single use or a combination thereof, rendering them to be inadequate in responding to the global health crisis for the underserved.

Addressing these unfulfilled needs, here we report here the first of its kind highly affordable, simple, accessible, user-friendly, nucleic acid based rapid POC diagnostic test that can simultaneously detect multiple viruses or viral strains with a broad spectrum of deployment ranging from hospitals and clinics to underserved community settings within 45 minutes. The test operates on a generic RT-LAMP-based assay implemented in a pre-programmable POC device that needs no particular alteration for different gene-specific targets. Unlike other reported assays for such purposes that either mostly leverage single-use fluidic cartridges driven by external fields for automated liquid handling operations or have been implemented mostly in controlled environment of a very lower number of cohorts to fulfilment of clinical validation, the present testing method deploys a one-for-all integrated thermal-cum-optical analytic unit in a closed community-based cohorts’ with execution of RT-PCR and RAT side by side. Further, our tests are executed therein within hermetically sealed standard PCR tubes, connecting the entire pipeline of sample to answer with only one-time dispensing of a reaction mastermix that ensures time-synchronized execution of the concomitant reaction steps without intermediate manual intervention. This robust operative embodiment completely eliminates the need of specialized custom-equipment such as any complex disposable cartridge and sophisticated fluorescent reader that can otherwise act as an obstacle to the affordability and deploy ability of the assay. A custom-made mobile application empowered by machine learning ensures accurate colorimetric detection, nullifying ambiguities due to visual dilemma or ambient effects. Further, the absence of downstream sample handling makes it suitable for use in a wide variety of settings without the need of any specialized operator. We demonstrate the efficacy of this test in the accurate differential diagnostics of influenza A and SARS-CoV-2 infection in an underserved rural setting with no specialized operator and laboratory control, reporting an overall outstanding clinical sensitivity of more than 96% and specificity more than 98% for field trials on 1500 cohorts, hence establishing itself as a “rapid nucleic acid test” of its kind.

## 2. Materials and Methods

### 2.1 Designing of RT-LAMP Primers for SARS-CoV-2 and Influenza A Detection

According to the principle of the RT-LAMP, one primer set contains three pairs of primers, specifically, inner primers pair: forward inner primer (FIP) and backward inner primer (BIP); outer primers pair: F3 primer and B3 primer; loop primer pair: loop primer forward (LF) and loop primer backward (BF), which can target six independent regions of the nucleotide sequence. In this study, two primer sets were designed against two different conserved target regions in the SARS-CoV-2 genomic RNA that are present within the nucleocapsid (N) gene (50) and the Spike (S) gene. For designing S gene targeting primer, the S gene encoded sequences of different variants of SARS-CoV-2 were derived from the NCBI Gen-Bank database. Nucleotide sequence of spike ORF of different SARS CoV2 variants of concern were aligned using Multiple Alignment Fast Fourier Transform (MAFFT) as a high-speed multiple sequence alignment program. Positional nucleotide summary was analyzed from the multiple alignment file in Bio-edit sequence editor. On the basis of conversation, a heat-map was generated (Fig. S1 of Supplementary Information). The conserved first 200 nucleotides of spike ORF was selected as target for the primer designing. Then six sets of primers were designed on the basis of these sequences for RT-LAMP reaction using PrimerExplorer V5 software (Fujitsu Limited, Tokyo, Japan). Co-infection of influenza A was tested targeting the M gene of influenza genome. The primers for RT-LAMP targeting M gene were designed similar to above. All primers (Integrated DNA Technologies, India), obtained in lyophilized form, were dissolved in Dnase, Rnase free ultrapure water (Invitrogen, Thermo Fisher Scientific) to a concentration of 100 µM and stored at -20 °C. 10X primer mix (1.6 µM FIP & BIP, 0.4 µM LF & BF and 0.2 µM F3 & B3) was prepared for use in the RT-LAMP assay.

### 2.2 Designing and Synthesis of Positive Control

Specific regions of the SARS-CoV-2 genome (Wuhan Hu-1) were synthesized for generating positive control, and purchased from Invitrogen. Each region contained a T7 RNA polymerase promoter in its 5’ end. The cassettes were cloned in pUC19 vector. Template for *in-vitro* transcription was prepared by PCR amplification of the cassette. The PCR purified DNA fragment was used as template for *in-vitro* transcription. T7 RNA polymerase (Roche, USA) was used to synthesize the positive control RNA. The *in-vitro* transcribed RNA was purified using Trizol reagent and its quality was checked by NanoDrop One Microvolume UV-Vis spectrophotometer (Thermo Scientific) and Urea PAGE (Figure not provided).

Influenza A/H1N1/WSN/1933 viruses (lab strain) were amplified in MDCK cells. Cell culture supernatant was collected and clarified by centrifugation and viral titre was determined by Plaque assay. The supernatant was diluted in Phosphate buffered saline (PBS) to prepare dilutions containing 10^6^ to 10 plaque forming units of virus per micro litre. These dilutions were used for the positive control for influenza A virus detection.

### 2.3 Optimization of the RT-LAMP Reaction System

Initially, short *in vitro* transcribed (IVT) RNA fragments of SARS-CoV-2 genome were utilized here as the positive control to optimize the RT-LAMP assay. The RT-LAMP reaction system (10 µL of volume) included with 5 µL WarmStart Colorimetric LAMP 2X Master Mix (New England Biolabs, USA, Cat #M1804L), 1µL 10X primer mix and 4 µL template. The entire isothermal process including heating at 65 °C from 0 min to 60 min to amplify the template and 95 °C for 5 min to terminate the reaction, was performed in a designated portable device (Figure 1A). The time of amplification step was optimized at time gradients (0, 20, 30, 40, 50 and 60 min). For each reaction, at least three independent replicates were performed. Negative control (NC) was performed in every experiment by using nuclease-free water instead of the positive samples. Upon completion of the thermal protocol, the reaction tubes were removed from the heating block and cooled to room temperature. The results were analyzed by the naked eyes and imaged using a smartphone with default setting. In addition, the RT-LAMP product formation was confirmed by 1% agarose gel electrophoresis. The same strategy was taken to optimize the test for the detection of influenza A virus using the prepared positive control samples of various dilutions containing 10^6^ to 10 plaque forming units of influenza A virus per micro litre.

**Fig. 1.**
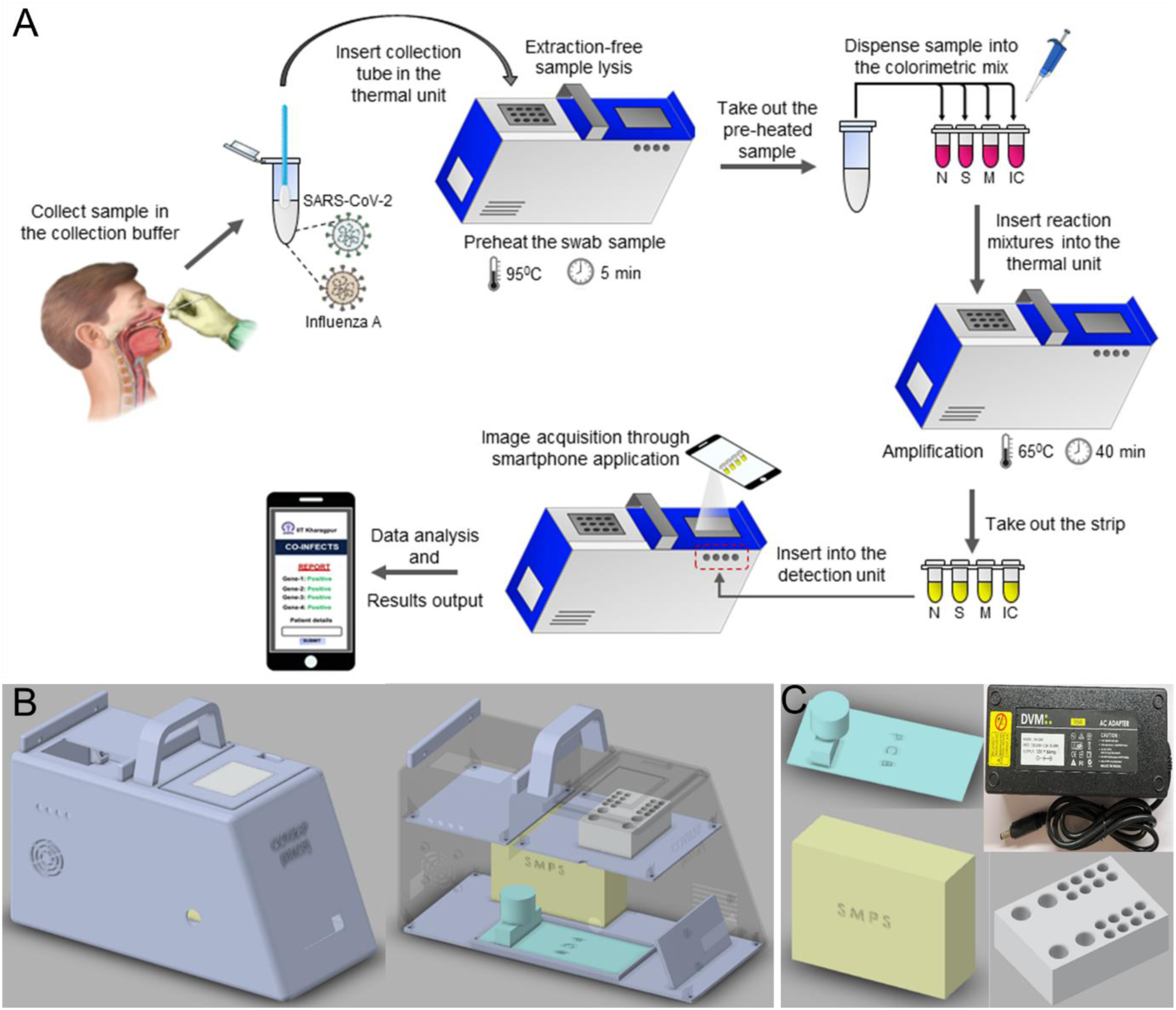
(A) Overview procedure for the sample to result POC detection of SARS-CoV-2 or influenza A viruses infection or co-infection by both. The one-for-all device technology for simultaneous detection of the test genes for multiple infections was designed using 3D modelling techniques (B) The assembled device showing outer view (left side), including the reaction space with sliding window, display and photo capturing portions. The device is light weight (<500 g) and can be carried by hand to disseminate the diagnostic test virtually anywhere. The open section view (right side) shows the internal features of the reaction and the imaging modules. The microcontroller operated heating unit conducts the nucleic acid amplification assay in the reaction tubes where the reaction mastermixes are dispensed into the inactivated test samples. (C) The modular device houses microcontroller, power adapter, heating block, SMPS and PCB units as shown separately.

### 2.4 Analytic Stability, Diagnostic Sensitivity and Specificity

To evaluate the stability of the test reaction kits, the colorimetric master mix and primers were premixed and placed in different reaction tubes in different sets and kept at -20 °C. Every 24 hours post incubation, fixed copies/dilutions of positive control virus were added to the tubes and reaction was performed. The stability was investigated up to 7 days. The control test was also performed with a freshly prepared reaction mix. Besides, real-time fluorometric assessment of reagent stability was also investigated by storing the mastermixes for up to 72 h followed by conducting reaction in every 24 hours and by storing the mastermixes at different temperature conditions (4 °C, -20 °C and room temperature) followed by assessing the reaction efficacy.

The gene wise (SARS-CoV-2 specific) sensitivity of colorimetric RT-LAMP was evaluated using various concentrations of the standard influenza A virus test RNA fragments (5 copies to 1 × 10^7^ copies per microliter). 4 µL of RNA template of each gene specific standard was added to the 10 µL of volume of the LAMP reaction. The reaction mixture was incubated at 65 °C over the pre-set time duration. At, the end of the reaction, the color difference between positive and negative samples was analyzed. Each experiment was repeated thrice. Besides, M gene specific sensitivity of influenza A virus detecting RT-LAMP protocol was evaluated using different dilutions containing 10^6^ to 10 plaque forming units of the virus per micro litre. The dilutions were heat inactivated for 5 minutes at 95⁰ C. 4 µL of the heat inactivated sample was used to determine the sensitivity of the assay. This assay was also repeated thrice.

Specificity of the S gene specific primer was evaluated against different variants of SARS-CoV-2 viz. alpha, kappa, delta and omicron having different Ct values. RNA was extracted from these strains and RT-qPCR was performed to quantitate the number genome equivalent present per microliter of the resuspended solution. Subsequently, 1×10^4^ copies/µL of each RNA sample (4 µL) were subjected to 6 µL of volume reaction mixture and possible visual color change was observed. The specificity of M gene target was also evaluated against different variants of influenza A virus viz. H1N1/WSN/1933, H1N1/PR/8/1934, H3N2 & H1N1 A/California/07/2009.

Gene wise (N, S & M) quantitative specificity of RT-LAMP was also compared with rapid antigen test (RAT) results. Herein, gene wise quantitative specificity of RT-LAMP assay was ascertained against 1500 clinical samples depending on the Ct value obtained from the gold-standard RT-qPCR. On the basis of sensitivity and specificity of the *in-vitro* test, a Receiver Operating Characteristic (ROC) was also constructed to estimate the on-field efficacy.

### 2.5 Clinical Samples and Ethics Statement

Oro-nasopharyngeal swab samples were collected in phosphate buffered saline (PBS) solution from patients/volunteers with their informed consent as per the approval of the Institutional Ethical Committee based on the declaration of Helsinki guidelines. Some amount of each swab samples was used for RNA isolation using HiPurA™ Viral RNA Purification Kit (Himedia India) according to the manufacturer’s protocol and the remaining amount was stored for direct sample to result testing purpose.

### 2.6 Co-infection Sample Constitution and Detection of Multiple Respiratory Viruses

Both SARS-CoV-2 positive and negative human nasal/oral swab samples were spiked with influenza A virus purified from the cell culture supernatant. 100 plaque forming units of influenza A/H1N1/WSN/1933 were mixed with 20 μL of clinical samples collected from SARS-CoV-2 infected and non-infected healthy human volunteers. The spiked samples were heat inactivated for 5 minutes at 95 °C. 4μL of the heat inactivated samples were used to detect both SARS-CoV-2 and influenza A virus co-infection.

### 2.7 System Design and Connectivity for Deployment in Underserved Community Settings

The main motive of this study was to establish the efficacy of a POC nucleic acid test in detecting respiratory virus infection caused by SARS-CoV-2 and/ or influenza A viruses concurrently, as an illustrative example of its multiplex capabilities. We assessed the sensitivity and specificity of each set of primers targeting individual genes of SARS-CoV-2 and influenza A viruses when used in a multigene format. The study was continued (June 2021 to December 2021) until sufficient human samples (more than a thousand) had been investigated to get a conclusive outcome of the on-field test performance in uncontrolled settings. Finally, we investigated the efficacy of this test in screening the co-infection caused by SARS-CoV-2 and influenza A viruses. We developed a system that can be easily deployed for the high-throughput respiratory diagnostics. The system included with technology hub where the testing kits and POC devices were manufactured. The system also included with a central distribution point for test kits to handout in defined quantities to workgroups on the campus for the detection of respiratory infections of suspected individuals who came at the PHC where sample collection, processing, and detection process were performed by designated workgroups. The participants’ details viz. name, contact number, age, vaccination status, symptoms and co-morbidities were well-documented. At the same time RAT were also performed side by side for cross-verification. The collected clinical samples were also shifted to the central distribution point for storage purpose. RNA from these clinical samples were extracted at technology hub and then distributed along with RT-PCR kits through central distribution point to the multispecialty hospital for the cross-verification by RT-PCR test. The RT-PCR test was used here as a gold standard test. Then, the datasheet containing the results, clinical parameters of patients, were send to analysis hub for further gene wise analysis of the data, co-relation with clinical parameters, and upload of the result in cloud database.

### 2.8 Field Dissemination and Testing

Community based testing of the RNA-extraction-free single-step swab-to-result method was conducted over a period of seven months at a rural primary health care centre (B.C. Roy Technology Hospital) located within the Campus of Indian Institute of Technology Kharagpur. The performance of RT-LAMP assay for SARS-CoV-2 and influenza A virus diagnosis was evaluated for 1500 human subjects. Swab samples, collected from the suspected individuals, were preheated at 95 °C for 5 minutes, and directly used for the assay without needing any formal extraction step. The whole procedure was performed by non-specialist operators outside any controlled lab using POC device. The positive samples were marked by visual color change in RT-LAMP detection which were detected through our smartphone application. Post the thermal reactions, the reaction tubes were inserted into the device’s specified slots to capture images of the tube contents using the smartphone camera, without needing to open the tubes up. A smartphone could be placed in a predefined position on the clear window of the device for the said purpose. The camera settings on the smartphone were hardcoded so that the user could not alter the same unwarrantedly. Overall procedure of detecting viral infection has been depicted in Fig. 1A. For declaring the test results, positive and negative tests were confirmed when the outcome was positive and negative for all of these target genes, respectively. Otherwise, the test result was concluded as negative. Consistency of the results obtained from the both tests was compared and the inconsistent test results were also further investigated considering the clinical and epidemiological findings of the volunteers viz. symptoms, vaccination status, age, co-morbidity etc. In parallel, the rapid antigen test (RAT) and/or RT-qPCR was performed with the same samples for the cross-verification of the test. RAT and RT-qPCR were performed using Indian Council for Medical Research (ICMR) approved test kits. Finally, the sensitivity, specificity, false/true positive, false/true negative, accuracy, were calculated and compared with RT-qPCR.

### 2.9 RT–qPCR Benchmarking

RT-qPCR was performed using multiplex RT-PCR Kit (ICMR approved) in parallel for the detection of SARS-CoV-2 specific RNA and influenza A virus specific RNA according to the manufacturer’s protocol. The performance of the RT-qPCR reaction was validated every time using a positive control supplied with the kit. The positive samples detected through our test where cross-verified using RT-qPCR assay as a gold standard. The cycle threshold (Ct) values, obtained from the RT-qPCR, were compared with the results of present test in order to obtain a quantitative estimate of the efficacy of our testing procedure. The RT-qPCR result was considered positive when the Ct values of target genes were ≤35, and negative when the Ct values were >35 which in accordance with the RT-PCR kit manufacturer’s guideline.

### 2.10 Bioinformatics Analysis

For designing of S gene targeted primer of SARS-CoV-2, multiple sequence alignment was performed with the more than 1500 whole-genome sequences of SARS-CoV-2 using the MAFFT server (https://mafft.cbrc.jp/alignment/server/) (51). Then, the Bio-Edit multiple sequence editor (www.mbio.ncsu.edu/bioedit/bioedit.html) (52) was used to analyse the conservation of the whole genome (53). Conserved target regions corresponding to the particular gene were selected from the consensus sequence obtained from the multiple sequence alignment. The same strategy was adopted during designing of M gene specific primer of influenza A virus.

### 2.11 Statistical Analysis

Gene wise sensitivity (positive percent agreement) and specificity (negative percent agreement) were calculated as per the following:

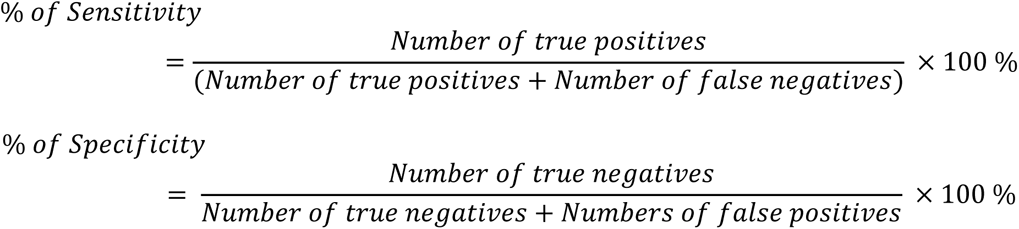

95% confidence interval (CI) was calculated for the data set using Clopper−Pearson′s method. Clinical data were analysed in Microsoft Excel (V 2013).

## 3. Results

### 3.1 Device Fabrication

The whole design and drawing of the instrument were performed using Autodesk Inventor Professional 2021. The device (Fig. 1B), developed in-house, is capable of executing the time-stamped isothermal protocols in automated sequence as synchronised by a microcontroller-controlled thermal unit. It was designed in such a way that all functionalities i.e. sample-to-detection processes could be integrated seamlessly. This was facilitated by an Arduino microcontroller, two aluminium heating cartridges, a DS18B20 one-wire temperature sensor, fan with heat sink assembly, optocoupler relay (DC) unit that may either trigger heating through cartridge heater or cooling through fan–heat sink assembly and a power unit as switchable alternative and another Switched Mode Power Supply (SMPS) unit. The respective modules of the device are shown in Fig.1C.

For executing our tests reported in this work, one thermal unit was pre-programmed at 95 °C for the lysis process, whereas the reaction unit was pre-programmed to operate at 65 °C. The thermal block features requisite numbers of slots to accommodate equivalent numbers of reaction tubes for several test reactions to run in parallel. The Arduino code, developed in-house, catered fetching the temperature data and executing the control functions in accordance (23). Post the lysis step, the reaction tubes, filled with the test reagents, were placed on the reaction unit. The heating control circuitry and power supply were operatively connected to the thermal block. The microcontroller code was in charge of gathering temperature data and carrying out three primary tasks: a) raising the temperature to the required target: in this function, the heater relay was prompted through an active low signal to an optocoupler. In the meantime, the cooling fan relay remained inactive; b) maintaining the temperature at the desired value: this function involved a temperature threshold to ensure that the temperature stayed within an acceptable tolerance, dT (typically 0.1 %). When the temperature fell below T - dT, the heating relay was activated, while exceeding T + dT activated the cooling relay. The temperature thus fluctuated between T + dT and T – dT, with the oscillations being minimized by the thermal mass of the heating block; c) lowering the temperature: similar to function (a), this function activated the cooling circuit instead of the heating circuit. Temperature readings were recorded every 3 s, and redundant checks were performed using temperature sensors to ensure accurate readings without any signal interference through the one-wire interface.

### 3.2 Smartphone Application for Image Analysis and Result Interpretation

A smartphone application empowered with machine learning, named CO-INFECTS, was developed in-house for the analysis of the colorimetric outputs of the thermal reaction and final interpretation of the test results (Fig. S2 of the Supplementary Information). CO-INFECTS was trained to read the colorimetric properties of the reaction tube contents and normalize the same with respect to pre-defined references, thereby making it easier to interpret the test findings without ambiguity. The captured image was resized by the application to process the image and generate test data. With its user-friendly interface and one-step operation requirements, this application aimed to deliver unambiguous and decisive test results. The RGB values of the reagent color and their change before and after the reaction were obtained with the help of OpenCV functions and then utilized as the observable measure to train the ML model in order to develop the image analytics technique. These were processed by in-built App functions automatically. In short, the captured images were resized through smartphone application and this application converted the image color channels from RGB to HSV for precise identification of the region of interest (ROI). The ML-model worked with the average value of R, G, and B following the exact selection of ROI for analysis and test outcome determination. In this case, Random decision forest classifier, an ensemble learning method for classification, was used. These image processing and feature extraction steps were done in both of the images captured before and after the isothermal amplification (Fig. S3 of the Supplementary Information).

### 3.3 Time to Detection, Analytical Sensitivity and End-point Result

The optimum amplification time and sensitivity to detect viral RNA was investigated for all of the primer sets. Time optimization of amplification step is shown in Fig. S4 of the Supplementary Information where it is observed that color changes from pink to yellow of the positive sample occurred within 40 minutes for all the target genes. Accordingly, in this study, 40 minutes was taken as the optimal duration for amplification. *In vitro* sensitivity assay was performed to determine the limit of detection (LoD) for each target gene. As shown in Fig 3A, the LoD for the SARS-CoV-2 N gene was 10 copies per microliter, whereas for the S gene it was 100 copies per microliter of the added RNA template. Below this resolution, the respective sets of primers showed false negative results. For the influenza A virus, the M gene target showed its LoD as 100 pfu/µL (Fig.2A). Below 100 copies, the sensitivity got compromised as anticipated for a simplified nucleic acid test executed without lab control.

**Fig. 2.**
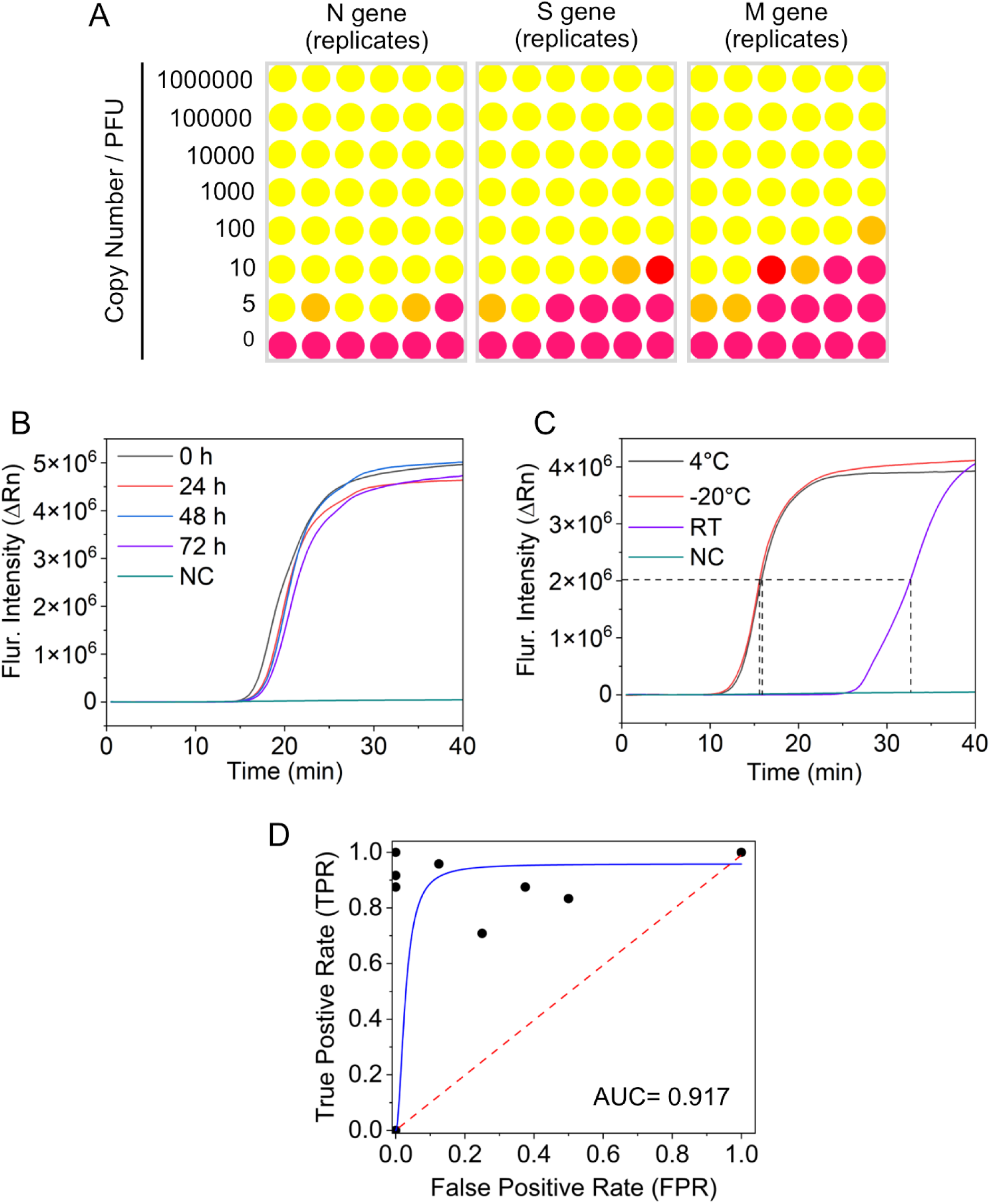
(A) Limit of detection of the method for individual gene targets (N and S) of SARS-CoV-2 detection. A known number of copies (ten-fold serial dilutions) of *in vitro* transcribed (IVT) viral RNA were amplified and detected by colorimetric RT-LAMP in sextuplets. Limit of detection of the RT-LAMP method for detection influenza A/H1N1/WSN/1933 virus targeting M gene. A known number of pfu/µL (dilutions containing 10^6^ to 10 pfu of virus per micro liter) of virus were amplified and detected by colorimetric RT-LAMP. The yellow colored circle represents positive detection, while pink colored and orange circle represents negative results and non-interpreted result for individual replicates. (B) Reagent stability characteristics in terms of time. Normalized real-time amplification curves of the master mixes stored at -20^0^C for up to 72 h. Different symbols indicate: 0 h of storage (black line), 24 h (red line), 48 h (blue line), and 72 h (purple line). (C) Reagent stability characteristics for estimating the optimal storage temperature conditions. Average threshold time as a measure of the test reaction kinetics and quantified in terms of the time for attaining 50% of the maximum fluorescent intensity for different storage conditions: 4 °C (black line), −20 °C (red line), and room temperature (purple line). NC stands for negative control. (D) Receiver operating characteristics (ROC) for the in-vitro assay. Blue solid line indicates the test and the 45° diagonal line (red dotted line) serves as the reference line.

**Fig. 3.**
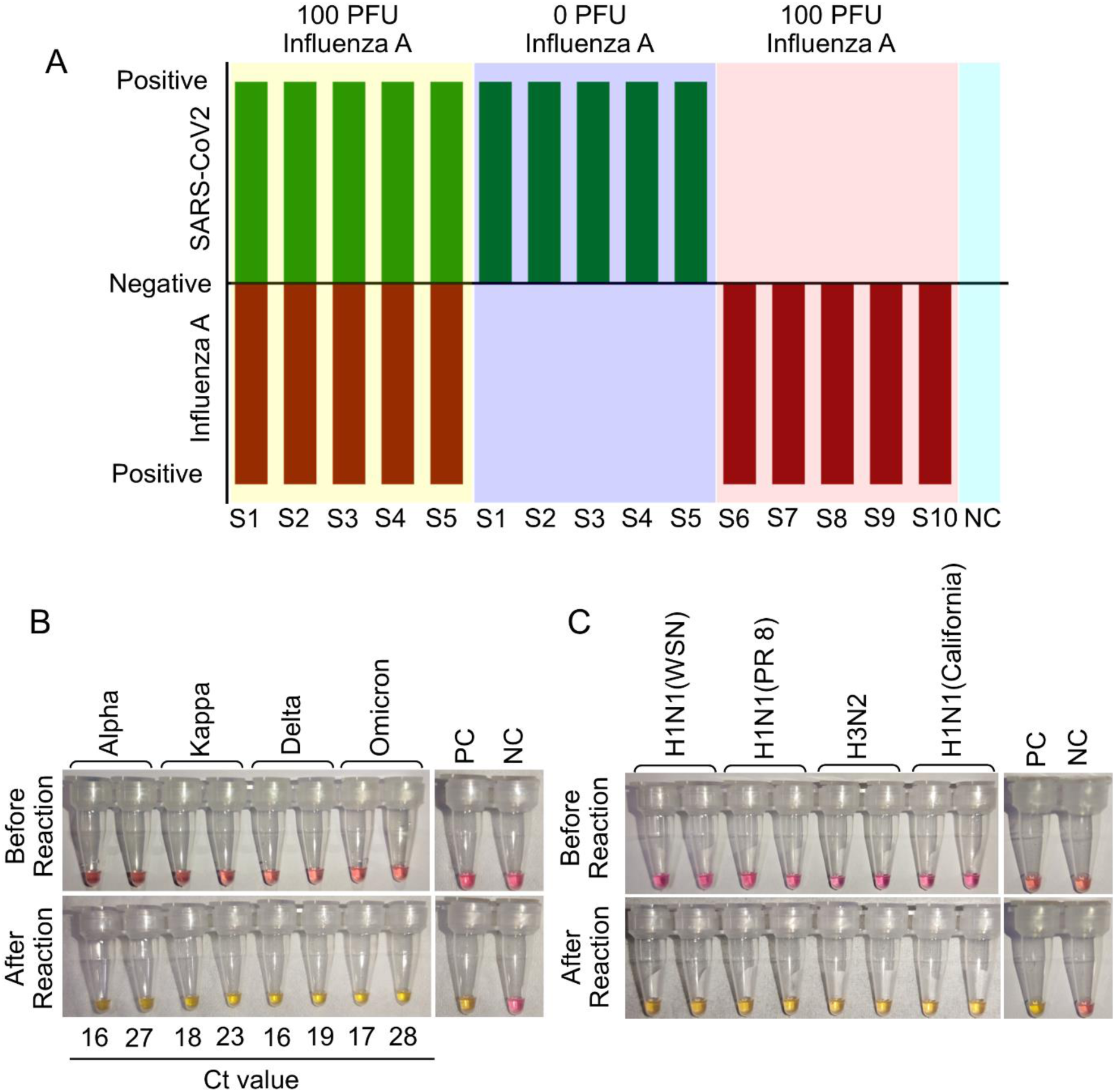
(A) Detection of co-infection using clinical SARS-CoV-2 positive samples having different Ct value and negative samples spiked with 100 pfu of influenza A/H1N1/WSN/1933 virus. S1 to S5 are the SARS-CoV2 positive clinical samples having different Ct values (S1- Ct 15/15; S2- Ct 15/17; S3- Ct 22/23; S4- Ct 16/17; S5- 30/29). Ct values are provided based on the N and E genes respectively. S6 to S10 are the SARS-CoV2 negative clinical samples. NC stands for negative control. (B) Specificity of the test towards the detection different strains of SARS CoV 2. (C) Specificity of the test towards the detection different variants of influenza A virus. Yellow color was visible for successful reaction. PC and NC indicate positive and negative control respectively.

### 3.4 Reagent Stability

In order to assess the reagent stability, we pre-assembled the probes and enzymes alongside the primers and the other reagents in the reaction tubes as ready-to-use mastermixes. Subsequently, these kits were stored at -20 °C for up to 72 hours before performing fluorometric isothermal tests. No significant change in the time threshold was observed for the stored reagents (Fig.2B). Additionally, the tests evidenced that storing pre-mixed test kits at -20 °C or at 4 °C for up to 72 hours did not alter the amplification profiles (Fig. 2C). This enabled us to supply the kits from a centralized -20 °C storage to decentralized normal refrigerators and keep them at a stretch for up to 3 days without performance compromise. In the central -20 °C storage, keeping the kits for 7 days at a stretch and performing amplification reaction with *in vitro* transcribed RNA at every 24 hours’ time interval evidenced uncompromised reactivity of the test reagents (Fig. S5 of the Supplementary Information).

### 3.5 Receiver Operating Characteristics

To further analyse the test performance, the Receiver Operating Characteristic (ROC) curve (Fig. 2D) was plotted on the basis of different concentrations of *in-vitro* RNA samples. The ROC illustrates the diagnostic ability of a binary classifier system as its discrimination threshold is varied, displaying the trade-off between sensitivity (true positive rate) and specificity (true negative rate) across different threshold values. A higher area under the curve (AUC) indicates better overall performance of the classifier in distinguishing between positive and negative cases. Typically, an AUC value of 0.5 indicates a lack of discrimination (meaning the test does not effectively distinguish between patients with and without the disease or condition). An AUC between 0.7 and 0.8 deems acceptable, while 0.8 to 0.9 is classified as excellent, and anything above 0.9 is considered outstanding. Positioning of the curve close to the top-left corner of the window ensured that the AUC was greater than 0.9, exemplifying highly favourable trade-off between the sensitivity and specificity at various decision thresholds (54,55).

### 3.6 Efficacy in Performing Extraction-free Testing in the Portable Device

For effective field-implementation in a swab-to-result format, it is essential to assess the compatibility of our test towards different swab collection modes. Generally, virus transport media (VTM) is recommended for storing or collecting pharyngeal swab samples (56). Hence, swab samples were collected from infection positive donor (Ct values ranges from 20-30) either in VTM or in phosphate buffer saline (PBS) and tested them using the pre-standardized RT-LAMP reactions targeted against three genes. Interestingly, sample collected in VTM showed no color change during RT-LAMP amplification, although agarose gel electrophoresis confirmed the amplification (Fig. S6 upper panel of the Supplementary Information). On the other hand, sample collected in PBS showed color change as well as amplification in agarose gel electrophoresis, suggesting that VTM is likely to interfere with the color change of the colorimetric RT-LAMP mastermix (Fig. S6 upper panel of the Supplementary Information). It is possible that the buffer capacity of the VTM resists the pH change of the RT-LAMP reaction mixture, thereby interfering with the changing of the color of pH indicator from pink to yellow. Encouragingly, tests performed directly with the swab samples collected in PBS gave identical colorimetric output to the test performed with RNA extracted from the same samples (Fig. S6 upper panel of the Supplementary Information). This confirmed that our reagents and method hold the ability to detect the infection directly from swab samples without prior extraction of RNA, which makes it suitable to be implemented at the extreme POC format.

Next, we used swab samples isolated from infected positive donors (both SARS-CoV-2 and Influenza-A) in PBS, which were then processed independently in a thermal cycler and our device to corroborate their performances. We showed that our device, with pre-programmed heating cycle (95 °C for 3 minutes, 65 °C for 40 minutes and 95 °C for 5 minutes) resulted in similar results comparable to the thermal cycler (Fig. S7 of the Supplementary Information) hence establishing a low cost, robust but accurate alternative of the thermal cycler or other similar expensive instruments for conducting the isothermal nucleic acid testing for any respiratory pathogen infections. In addition, the App-based result interpretation (Fig.S2 of the Supplementary Information) rendered this device ideal for implementation at the POC format by minimally trained and non-specialized personnel.

### 3.7 Multiplexed Detection of SARS-CoV-2 and Influenza A

Although the influenza A H1N1 is reported to be the most commonly detected amongst the SARS-CoV-2 co-infecting viruses found in patients (9,57), their differential diagnostic technologies for POC-based testing remained to be scarce. Towards assessing the efficacy of our technology in fulfilling these needs, we first used constituted clinical samples by spiking in 100 pfu of cultured, purified influenza A virus into the different clinical samples that have either positive or negative for SARS-CoV-2 infection with different viral loads (Ct values provided in Table S1) as quantified using RT-qPCR. Our test was successful in detecting and distinguishing influenza AA and SARS-CoV-2 infections directly from these samples (Fig.3A; Fig S8 of the Supplementary Information), which gave the necessary foundation for on-field testing and deployment.

### 3.8 Specificity in differential diagnostics of SARS-CoV-2 and Influenza A viral strains

Our field study was initiated during June 2021, during which different strains of the SARS-coV-2 virus were circulating in different parts of the world. In India, emergence of the highly pathogenic Kappa and Delta variants resulted in the devastating second wave of the pandemic around that time, with a spike in the number of daily cases and associated mortality rates. In order to ensure the success of our test on the field, we probed first the broader cross reactivity of our test against different variants of SARS-CoV-2. Clinical samples positive for infection with different SARS-CoV-2 variants (verified by RNA sequencing using COVID-Seq kits - Illumina at NIBMG, Kalyani, West Bengal, India) were collected from one of the medical institutions participating in this. The test successfully detected each of the variants of SARS-CoV-2 in clinical samples, even from the ones having low viral load (Fig.3B). Besides, the test was also capable of detecting two different laboratory strains and one pandemic strain of influenza A/H1N1 and a laboratory strain of influenza A/H3N2, thus substantiating its ability to diagnose most of the influenza A viruses circulating in human population during the seasonal outbreaks (Fig.3C). Together all these data confirmed the robustness of our test to be implemented in the field for the detection of the SARS-CoV-2 and influenza A virus associated respiratory infections, at the POC setting.

### 3.9 Field Dissemination at the Community Settings Unveiled Clinical and Epidemiological Patterns Associated with SARS-CoV-2 and Influenza A Virus Co-circulation

We implemented our test in a POC format at the primary health-care (PHC) facility located on-campus that serves the students, employees, large numbers of regular and occasional visitors and a limited neighbourhood population (Fig. 4). This centre does not harbour any sophisticated molecular diagnostic facility. Hence, all of the potentially-infected clinical samples collected herein are routinely transferred to other multi-speciality hospitals for RT-PCR test. As the COVID-19 cases peaked during the second wave of pandemic (March to October 2021), we engaged with a total of 1500 patients showing symptoms of respiratory illness and consented to voluntarily participate in the field trial-based evaluation of our test. Nasal and oral swab samples were collected from these individuals for detecting SARS-CoV-2 and influenza A virus infections via nucleic acid testing and also for the RAT specifically targeted towards SARS-CoV-2 detection at the point of collection as a screening measure as per regulatory guidelines. The same samples were transported and subjected to RNA extraction and RT-qPCR for the detection SARS-CoV-2 and influenza A virus infection as per the standard procedure in the established Hospital settings of the clinical partners of this trial.

**Fig. 4.**
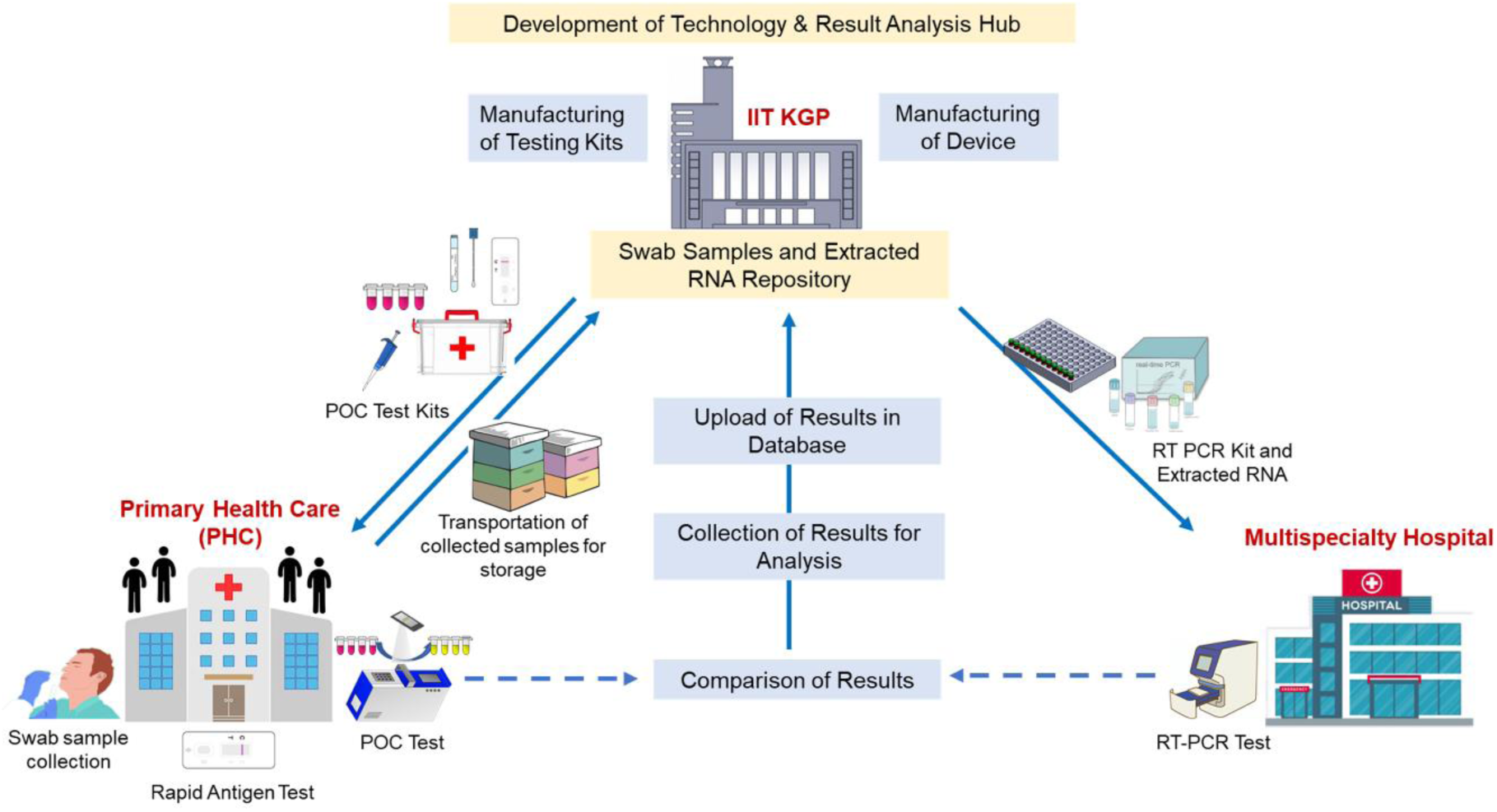
Concept design and system integration for a field trial-based validation at the underserved community settings for the detection of SARS-CoV-2 and influenza A virus infection. The testing kits and POC devices were manufactured at the technological hub from where the test kits and POC devices were distributed to the primary health care centre (PHC) in order to detect respiratory infections of suspected patients. Sample collection, processing, and detection process were performed at the point of care. Additionally, RAT was carried out side by side for cross-verification. RT-PCR test was also conducted using the extracted RNA of suspected individuals at multispecialty hospital. All test results were compared, analysed and upload to the database at analysis hub and finally informed the participants.

In our study period during June to December 2021, we found that daily cases of SASR-CoV-2 infection peaked during June to August followed by a steady decline during September and October and then again started increasing from November onwards. It should be noted that in India, the second wave of the COVID-19 pandemic was primarily caused by the Delta variant (B.1.1.617.2) spanned over the months of March to October 2021, followed by initiation of the omicron wave during December2021-January2022. Thus, our study conducted within the cohort was reflective of the overall dynamics of the COVID-19 pandemic nationwide. Interestingly, influenza A virus infections remained dormant during the initial phase of the study but started increasing during late October. In this period, a significant number of infections caused by influenza A virus and SARS-CoV2 occurred in parallel and as expected, resulted in a significant number of co-infections (Fig.5A). Among the four influenza types, i.e., Influenza A, B, C, and D, influenza A and B type infections commonly cause seasonal infections in humans. Although Influenza virus infections are reported throughout the year in India, during the winter season and the month of seasonal change, it surges by several fold (58,59). Accordingly, we divided our study period into two separate sub-periods for further analysis; one is pre-October during which SARS-CoV-2 single infections prevailed, and another is post-October when both SASR-CoV-2 and influenza A viruses jointly contributed to the respiratory infections within the community.

**Fig. 5.**
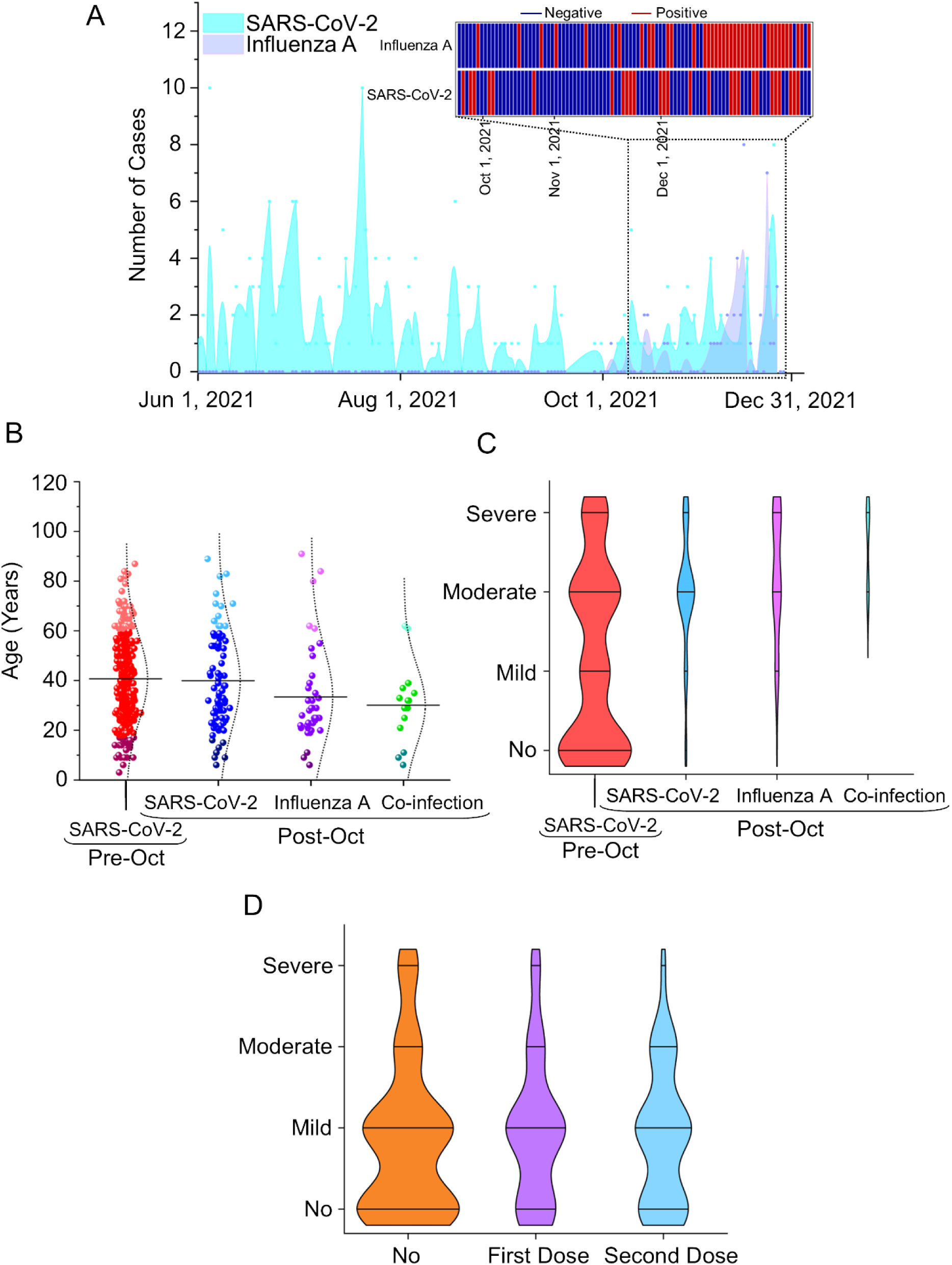
Clinical and epidemiological characteristics of study to detect SARS-CoV-2, Influenza A and their co-infection. (A) Timeline basis comparative analysis of infections caused by SARS-CoV-2 and influenza A. Inset: Showing the co-existence of both types of infections during flu season (post-October). (B) Age wise distribution of the infected persons by SARS-CoV-2, influenza A and by both. Maximum number of infected individuals were reported within the age group of 20-60 years in every case. (C) Overall distribution of degree of symptomatic pattern of infected persons depending on the source of infections. (D) Overall pattern of disease severity of infected participants depending on vaccination status.

A wide assemblage of symptoms was recorded among infection positive individuals. While the most common symptoms were cough, cold and fever; other symptoms like headache, tiredness, loss of taste or smell, sore throat were also reported, which is in line with the previous studies (60–62). Our analysis showed that both SARS-CoV-2 and influenza A virus infected a wide range of age groups starting from young citizens to old age people, over the entire period. However, maximum number of infected persons lied within the age group of 20-60 years with a median age of 40 year for SARS-CoV-2 infection and 35 year for influenza A infections (Fig. 5B).

Individuals who got co-infection accounted for 12.6 % (among positive cases) of the post-October study cohort. Mostly working age group, who were maximally exposed to the outer community, were found to be infection positive and reported to exhibit similar kinds of clinical symptoms (13) so that only symptom-based differential diagnostics was effectively impossible. In the pre-October cohort, significant percentage (59.6% out of 334 positive cases) of SARS-CoV-2 infected individuals showed mild to severe symptomatic pattern as well as remained asymptomatic. But, in post-October cohort, majority of the infection positive individuals (83.5% out of 79 positive cases) reported moderate to severe symptomatic pattern (Fig.5C). Importantly, co-infection by both the viruses lead to moderate or severe symptoms which may results from synergistic effects of the two pathogens in triggering the pro-inflammatory cytokines as described earlier (63–65). We found that the disease severity decreased with increase the vaccination doses (Fig.5D).

### 3.10 Multiplex Detection Efficacy for Testing Clinical Samples

Our testing was performed targeting N and S genes of SARS-CoV2 and M gene of influenza A and a comparative analysis between RT-qPCR (as gold standard test) and our test was conducted. Accordingly, the sensitivity and specificity of our test was determined. For the detection of SARS-CoV-2, the sensitivity and specificity of the test were found as 96.19 % (95% CI: 93.29-98.08) and 98.52 % (95% CI: 97.67-99.12) respectively (Fig.6). For the detection of influenza A virus, the sensitivity and specificity of the test were found as 95.75 % (95% CI: 92.50-97.88) and 97.63 % (95% CI: 96.46-98.57) respectively. Furthermore, the test’s sensitivity and specificity for identifying co-infection caused by both viruses were determined to be 95.17% (95% CI: 90.10-97.19) and 96.83% (95% CI: 95.76-97.77) respectively. This analysis confirmed that this single step swab-to-result test format integrated with our device makes this testing ideal to be implemented at the point of care format without compromising the detection rigor.

**Fig. 6.**
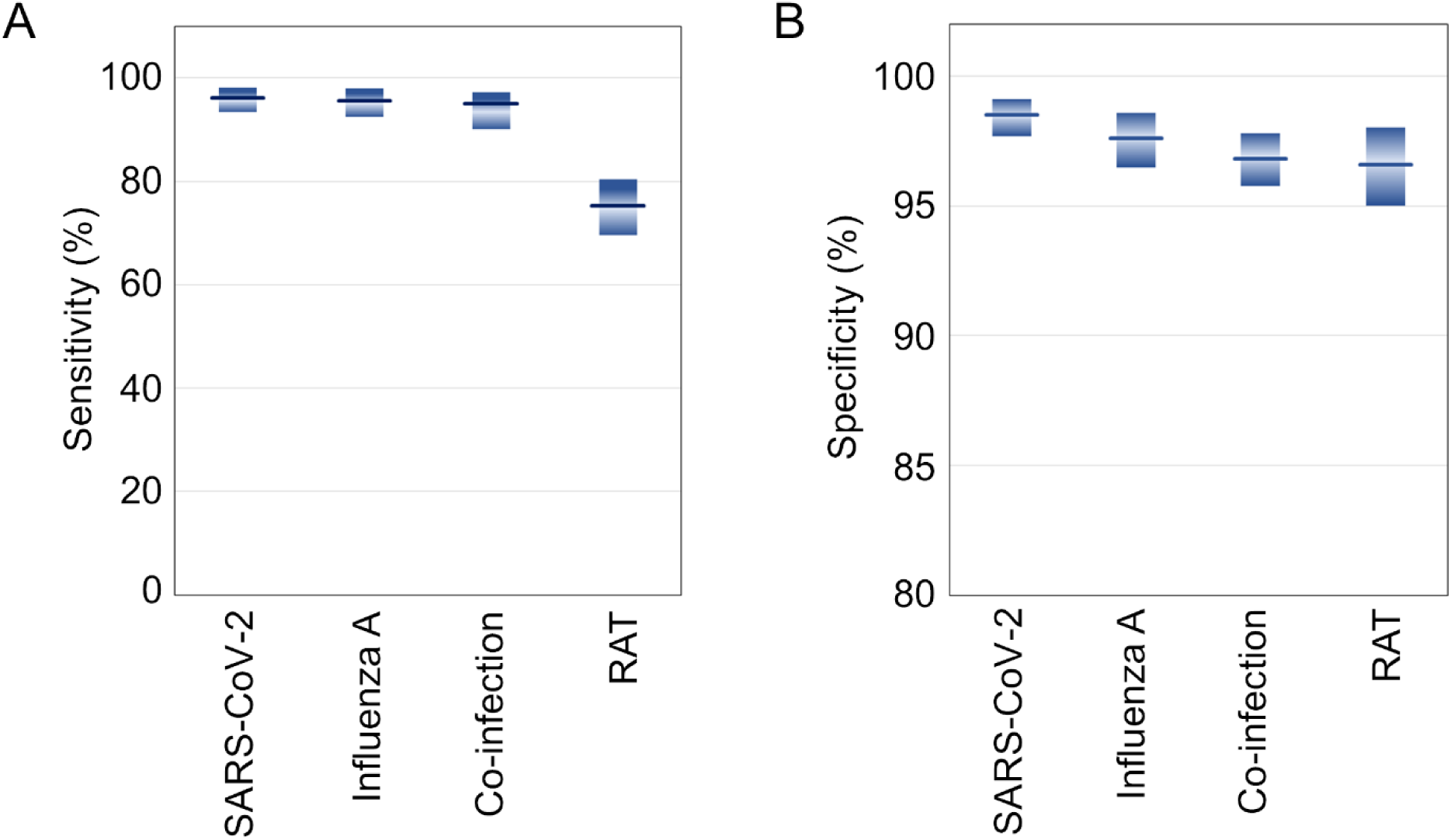
Box plots showing the comparative (A) sensitivity and (B) specificity of the test for the detection of SARS-CoV-2 infection, influenza A infections and co-infections from patient samples compared with rapid antigen test (RAT). The shaded bar represents the 95% confidence intervals calculated by Clopper−Pearson method

Additionally, rapid antigen test (RAT) was performed to compare the performance of our test with RAT, considering similar simplicity in the dissemination format. Since the RAT targets virus surface antigens that cannot be readily amplified unlike the target DNA in the nucleic acid tests, is known to suffer from lesser sensitivity and specificity (66). Our analysis confirmed that the sensitivity (75.28%) of RAT was much lower than the present test (> 90%), although the specificity was comparable. This established the value proposition and pitch-point of our test, exemplifying significantly higher sensitivity albeit similar user-friendliness and favourable cost proposition as compared to a common RAT test. It is also important to note, however, in the first place that RAT kits for multiplex respiratory co-infection detection do not exist in the common supply chain unlike the COVID-19 specific tests that hit the market in response to the pandemic challenge.

We also calculated the implementation costs acquired during the running phase of detection. The variable costs (reagents, consumables etc.), without fixed costs (rent of equipment, furniture, and laboratories) are summarized in Table 1. These cost estimates were based on the four-tube reactions for testing three viral genes (N, S & M) and one internal control (RNaseP). While the average cost of a RAT test is typically US $6.0, the average cost of our test was determined to be around US $3.0.

**Table 1.** Average cost per sample tested based on three viral genes and one internal control.

| Components | Cost/Reaction in USD |
| --- | --- |
| **With RNA Extraction** |  |
| Viral RNA Extraction | 0.54 |
| Colorimetric LAMP Mix | 1.76 |
| LAMP Primer Mix | 0.54 |
| **Total (INR)** | **2.85** |
| **Without RNA Extraction** |  |
| Colorimetric LAMP Mix | 1.76 |
| LAMP Primer Mix | 0.54 |
| **Total (INR)** | **2.31** |

## 4. Discussion

While the humanity is yet to be completely relieved from the devastation caused by the Novel Coronavirus Disease (COVID-19) pandemic, the World Health Organization (WHO) recently alerted on the upcoming “Disease X” which could be 20 times deadlier than the recent pandemic (67). As such unknown viruses threaten to unfold, multiplex testing facilities of their co-infection with other known infections of similar manifestation will continue to be progressively more imperative. Besides SARS-CoV-2, influenza A viruses are the major respiratory pathogens which has the history of causing five different pandemics in the last century, with the 1918-Spanish flu pandemic being the most devastating one. Apart from the pandemic outbreaks, influenza A and B viruses co-circulate during the flu season leading to high morbidity rates in specific age groups. In situations where influenza vaccination program is not strictly implemented, quick and repetitive diagnoses of high-risk populations and providing adequate medical facilities to the infected individuals would significantly reduce the impact of the such seasonal and occasional outbreaks. Co-occurrence of the SARS-CoV-2 infection further complicates this scenario. As could be predicted, SARS-CoV-2 and influenza viruses would frequently co-circulate and generate recurring epidemics (25,68). While our results verified this assertion, what appeared to be more concerning was the fact the co-infected patients showed moderate to severe symptoms as compared to the patients infected with either one of the viruses.

The present work is also likely to hold its unique implications from one-health considerations interlacing animals and humans, with particular concerns on an unprecedented recurring and recent occurrence of bird flu panzootic caused by High Pathogenic Avian Influenza (HPAI) H5N1 that already resulted in the deaths of millions of birds (69,70).Whereas a direct risk of humans being infected from such outbreak remains unknown, the same cannot be precluded for the personnel having to remain occupationally connected with the infected birds, including the wildlife researchers and poultry farmers. For such vulnerable, the zoonotic risk may get substantially elevated with potential mortality rates around 40 to 50% (71). Amidst the prevalence of other respiratory co-infections, the resulting impact on human health may compound to be lethal and thus requires widespread surveillance for prevention. This may call for the analysis of nasal and oral swabs or faecal samples in a seemingly healthy albeit vulnerable population of birds and humans. RAT and immunofluorescence assays, which are recommended for testing, have low to moderate sensitivity leading to higher false negatives compared with other molecular or RT-PCR tests or High Throughput Sequencing. Prior to genomic testing using RT-qPCR or High Throughput Sequencing for detecting the presence of influenza in a population with or without an accompanying co-infection, the present approach may thus provide an invaluable tool for large-scale screening by incorporating suitable primers in the test reaction mastermix.

The present multiplex framework is also likely to bear critical implications in targeting distinctive clinical management approaches without any speculation, empiricism or hit-and-miss trial, although, general and supportive care, especially for mild illness, is similar for both. For example, for COVID-19, nirmatrelvir-ritonavir is strongly recommended for patients at high-risk of severe disease, whereas oseltamivir is known to be effective for influenza, especially if administered within the first 48 hours after the onset of the illness. From this point of view, the present multiplex test also holds significant practical value for healthcare professionals who can selectively direct the protocols for infection-specific treatment. Given that various respiratory infections necessitate distinct treatment approaches, such POC respiratory multiplex tests should aid the physicians in delivering efficient therapies and reducing the risk of hospital-acquired infections. Furthermore, vulnerable co-morbid individuals may experience improved medical management as the test outcomes may favourably influence their disease treatment strategies. Reported evidences (72) already indicate that improved use of a class of antiviral medications, antibiotics as well as timely and judicious decision making on hospitalization and ICU admission may become feasible with accurate detection of the active presence of particular variants of acute respiratory infections or otherwise. Another encouraging approach that may emerge from the present work is a new means of allocating resources to adaptable diagnostic platforms with a versatile core platform technology, enabling the seamless replacement of pathogen probes to address emerging respiratory health threats via simple pre-programming of the test protocol that may run seamlessly in the device without the need of any intermediate manual processing.

## 5. Conclusions

We reported a highly affordable nucleic acid based POC test for detecting multiple respiratory infections via a RT-LAMP based platform technology, where the tests could be disseminated with the simplicity, user-friendliness and cost proposition similar to common rapid tests. Both SARS-CoV-2 and influenza A virus infections carried similar symptomatic pattern, as the COVID-19 pandemic continued into flu season. This made it difficult to distinguish SARS-CoV-2 and influenza A virus co-infection without proper laboratory confirmation. In this direction, we successfully used a single test to distinguish the SARS-CoV-2 and influenza A infections. The tests showed no cross-interference among the target genes for SARS-CoV-2 and influenza A virus, with acceptably high sensitivity and specificity when deployed on the field via randomized trials. These tests could be disseminated at on-campus community settings via a rural primary healthcare center with no requirement of laboratory control and specialized analysts. Our field trials not only exhibited the efficacy in differentially detecting the viral respiratory infections at different stages but also unleashed the implications of different epidemiological factors, thereby establishing a model for public health policy recommendation at identified geographical locations and cohorts depending the prevailing and circulating pathogenic strains. This approach holds tangible benefits for enhancing preparedness for future epidemics and pandemics, by assuming a crucial role in the surveillance and containment of potential disease outbreaks, where the target viral strains would need to be dynamically accommodated in the infection surveillance. Being a generic platform technology, this aspect could be readily implemented in our framework by simply altering the primer design for the said targets without needing any hardware or software alteration. This, in turn, holds the potential of ensuring fair and affordable access to testing for vulnerable communities to mitigate health disparities related to respiratory infections, without needing to wait for the access to high-end laboratory tests in time-critical scenarios. The useful implications thus appear to be much more far-reaching beyond the illustrative field work exemplified herein, with a vision of enhancing the global preparedness as future epidemics and pandemics unfold.

## Author Contributions

**Sudip Nag:** Primer and assay design, conceptualization, research planning, experimentation, data analysis, original manuscript writing, editing and revision.

**Saptarshi Banerjee:** Primer design, help in the development of *in-vitro* assay of influenza A detection, help in analysis of Fig.4 B, C and D data.

**Aditya Bandopadhyay:** Design and fabrication of POC device.

**Indranath Banerjee:** Sample collection, field validation and analysis

**Subhasis Jana:** Access of RT-PCR experimentation.

**Arindam Mondal:** Funding acquisition, ideation and concept development, research planning and supervision, manuscript writing and final editing.

**Suman Chakraborty:** Funding acquisition, ideation and concept development, research planning and supervision, manuscript writing and final editing.

## Acknowledgements

All authors acknowledge the Indian Institute of Technology Kharagpur and the Indian Council for Medical Research (ICMR) for facilitating different components of the work. S.C. acknowledges the DSIR, Government of India, for financial support through the CRTDH Project on Affordable Healthcare and DST (SERB), Government of India, for the Sir J. C. Bose National Fellowship. A.M. acknowledges ICMR (VIR/COVID-19/19/2021/ECD-I) and CRG-SERB (CRG/2022/003628; Date: 11.07.2023). S.C and A.M also profoundly acknowledge ICMR-DHR CoE project. S.N. acknowledges ICMR for providing Research Associate Fellowship (File No: 5/3/8/14/ITR-F/2022-ITR). The Authors acknowledge Ms. Subhanita Roy, Ms. Ranjini Chowdhury for their help and support (preparation of testing kits, help in RT-PCR test) during field validation and also Mr. Shyamal Manna, Mr. Sandip Maji for their help in swab samples collection and field testing. The Authors acknowledges Mr. Sohom Banerjee Mr. Ankan Bairagi, and Mr. Akasdeep Mondal and Dr. Sujoy Biswas for their support in device design, fabrication and other contributions related to the concerned technology development. The Authors also acknowledge Sreyansh and Srinivas for helping with the smartphone application development.

## Conflict of interest

The authors have no conflict of interests to declare on the reported work.

## Ethics Declarations

This study is approved by Indian Institute of Technology Kharagpur Biosafety and Ethics Committee obeying ethical principles for medical research involving human subjects according to the declaration of Helsinki guideline and obeying the guidelines of Indian Council for Medical Research (ICMR). In addition, for the investigation involving human subjects, informed consent has been obtained from the participant involved.

## Data Availability

The data supporting the findings of this study are available within the article and its Supplementary Information. The raw and analysed datasets generated during the study are available for research purposes from the corresponding authors on reasonable request.

## Notes

### Competing Interest Statement

The authors have declared no competing interest.

### Author Declarations

Institutional Ethical Committee, Indian Institute of Technology Kharagpur, India

