## Supplementary Information for "CO-INFECTS: A Highly Affordable, Portable, Nucleic-Acid-Based Rapid Detector of Active Respiratory Co-Infections via “Swab-to-Result” Integration"

**Table S1:** Details of SARS-CoV2 clinical samples

| **Sample Number** | **Sample Details** |
|---|---|
| S1 | SARS-CoV-2 (Ct 15/15) |
| S2 | SARS-CoV-2 (Ct 15/17) |
| S3 | SARS-CoV-2 (Ct 22/23) |
| S4 | SARS-CoV-2 (Ct 16/17) |
| S5 | SARS-CoV-2 (Ct 30/29) |
| S6 | Negative |
| S7 | Negative |
| S8 | Negative |
| S9 | Negative |
| S10 | Negative |

** Ct values are provided based on N and E genes.

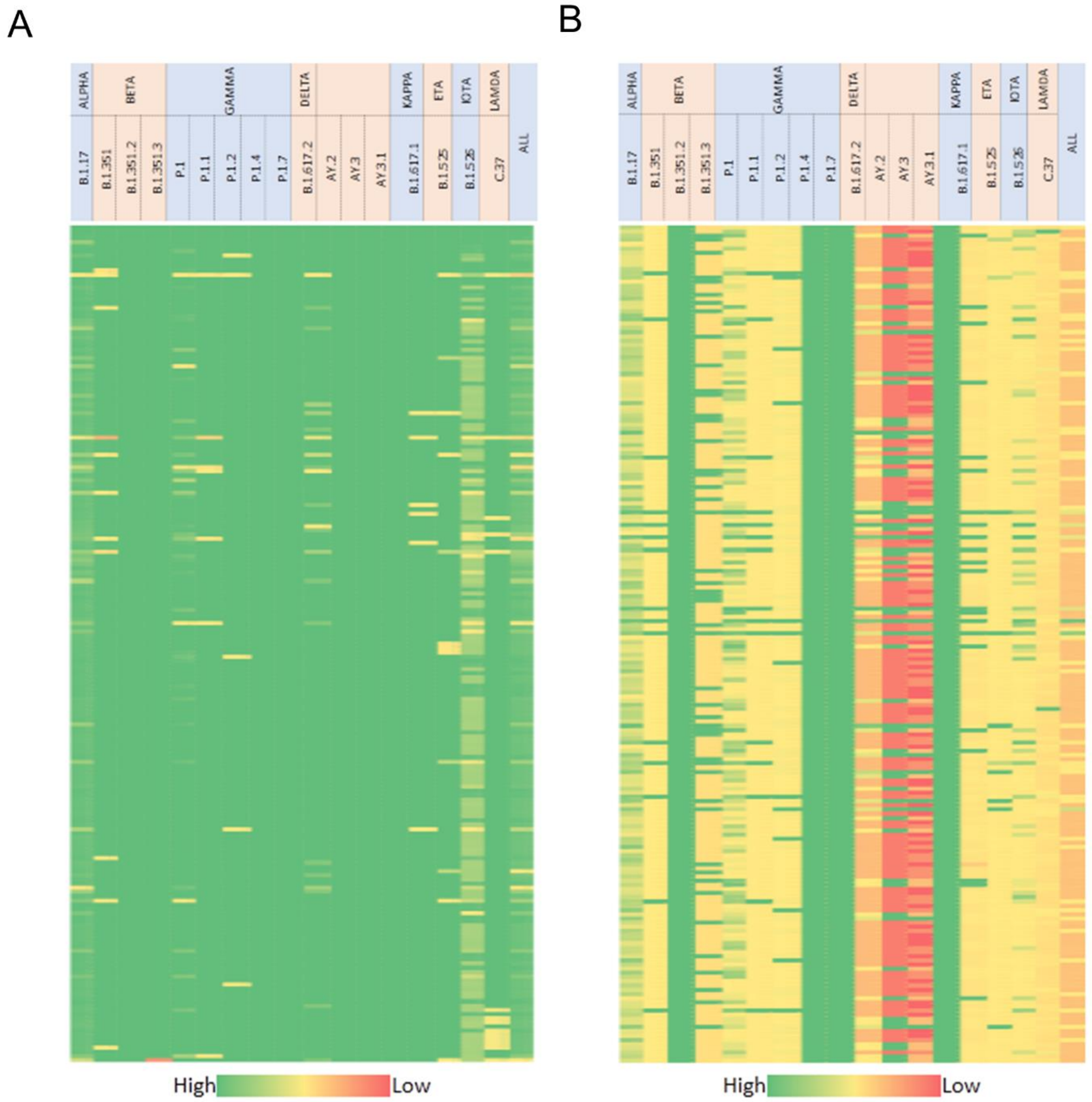


**Fig. S1**: Heat map showing conservation of nucleotide sequence for different variants. (a) 21563-21763 nucleotide, (b) 22556-22756 nucleotide region.

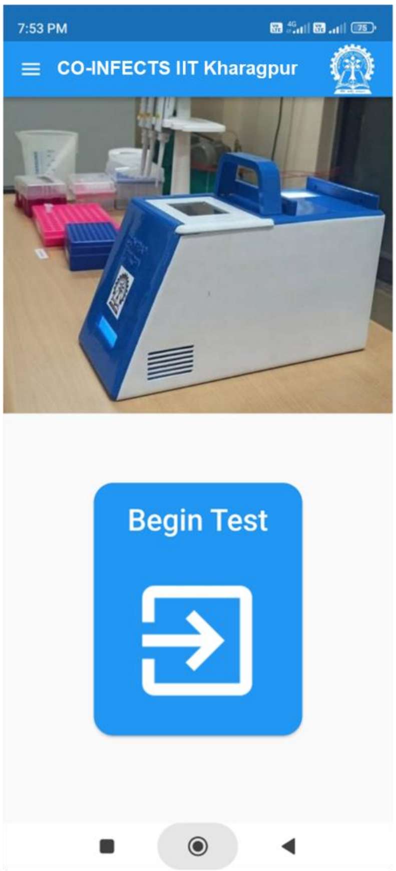


**Fig. S2:** Smartphone application (CO-INFECTS) for result analysis

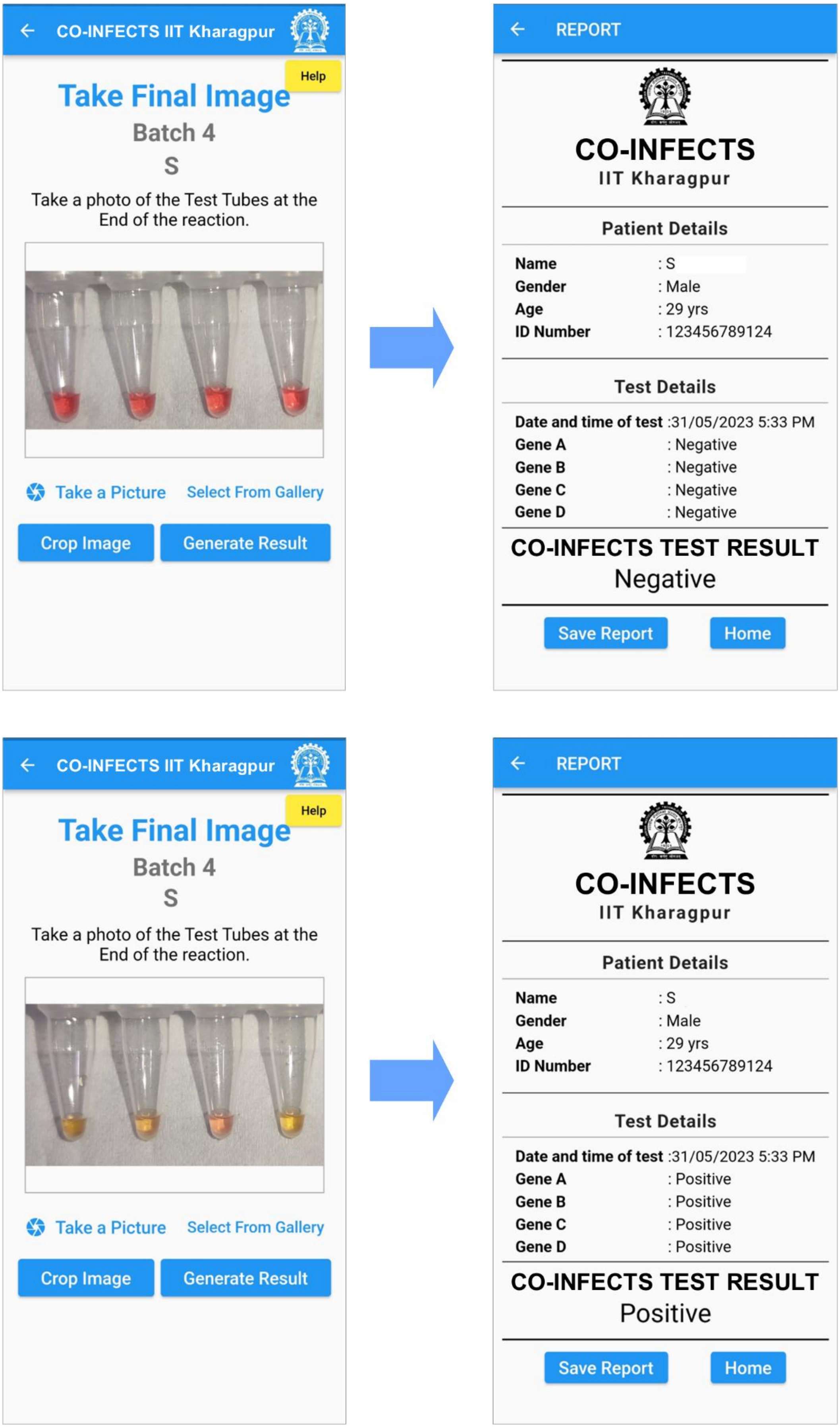


**Fig. S3:** App based interpretation of results obtained from RT-LAMP test

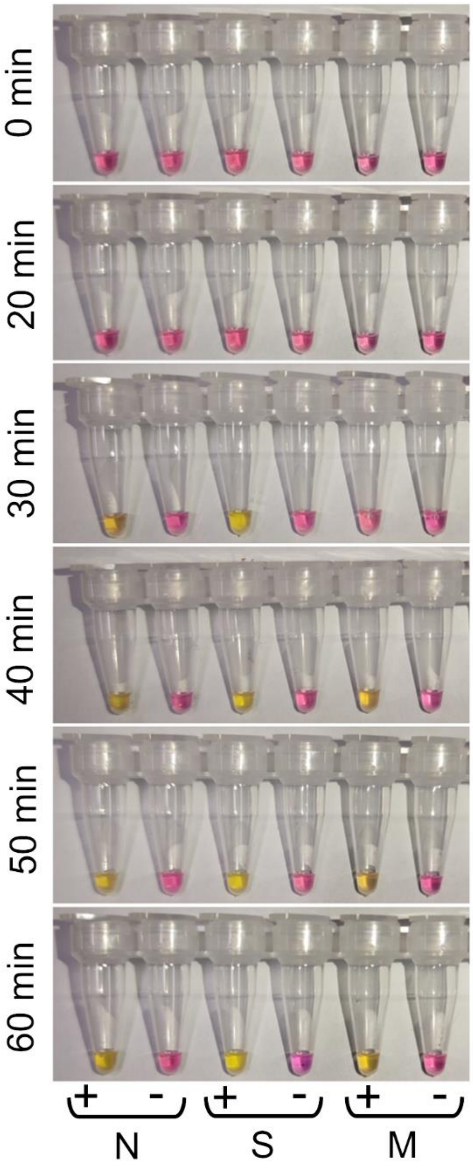


**Fig. S4:** Optimization of reaction time was performed at 65°C temperature on the basis of three set gene specific primer (N, S and M). Typical yellow color changes of all sets were observed within 40 min. N: Nucleocapsid gene, S: Spike gene, M: Membrane protein gene

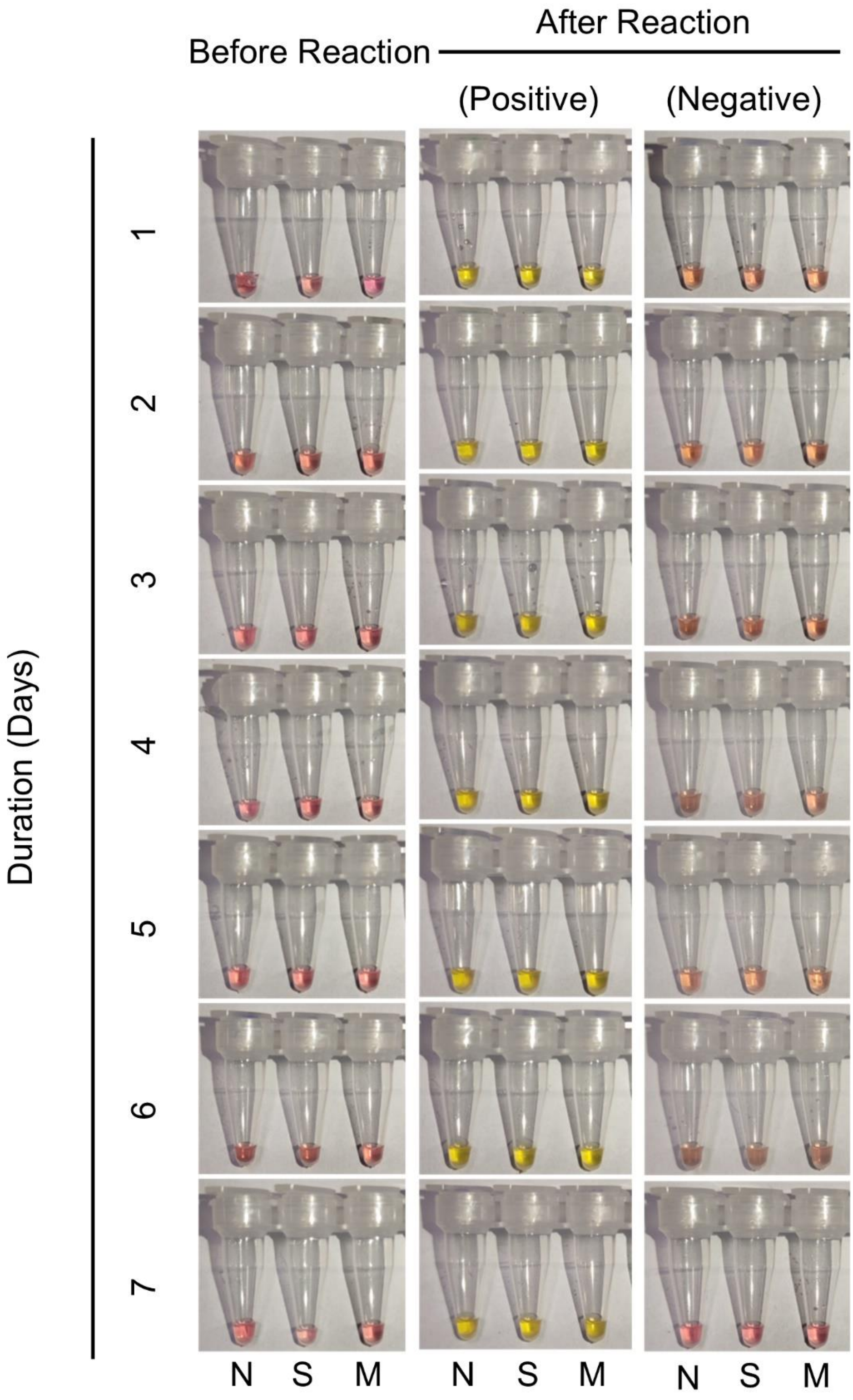


**Fig. S5:** Testing the stability of the reaction kit. The primers and enzyme master mix were prepared and stored at 4°C up to seven days. Every 24 h post incubation, positive control was added to each tube and performed RT-LAMP targeting the three genes (N, S, M). N: Nucleocapsid gene, S: Spike protein gene, M: Membrane protein gene

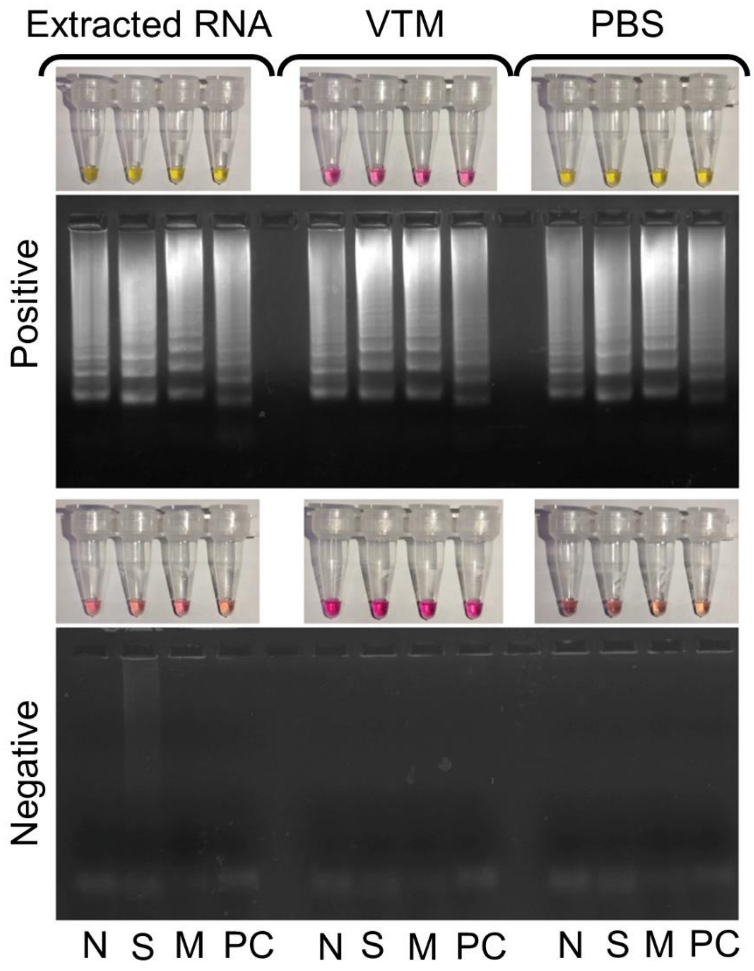


**Fig. S6:** Performance evaluation of test protocol on the basis of three targeted genes (N, S, M) using clinical sample collected in VTM and PBS compared to extracted RNA from the same sample. Positive sample showed typical ladder pattern of band of a successful RT-LAMP reaction along with color changes to yellow. Negative samples showed no obvious color changes as well as band pattern. N: Nucleocapsid gene, S: Spike gene, M: Membrane protein gene, PC: Positive control

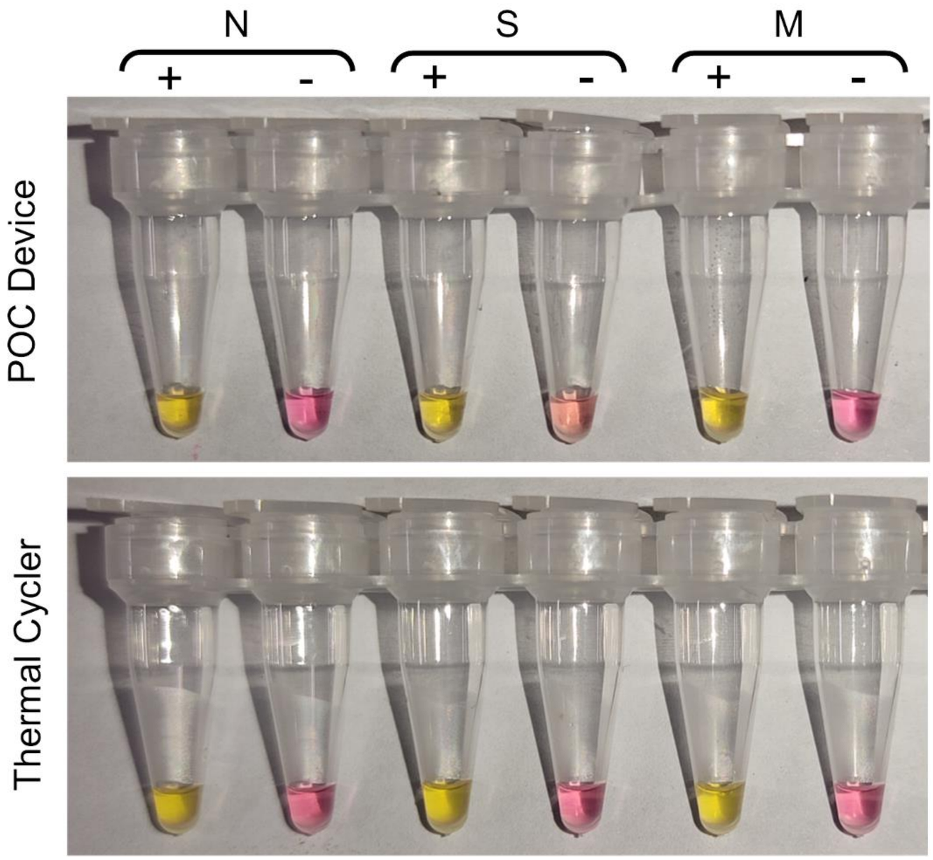


**Fig. S7:** RT-LAMP reaction performed in in our detector and thermal cycler to check the present test performance as compared to that using a thermal cycler. N: Nucleocapsid gene, S: Spike gene, M: Membrane protein gene

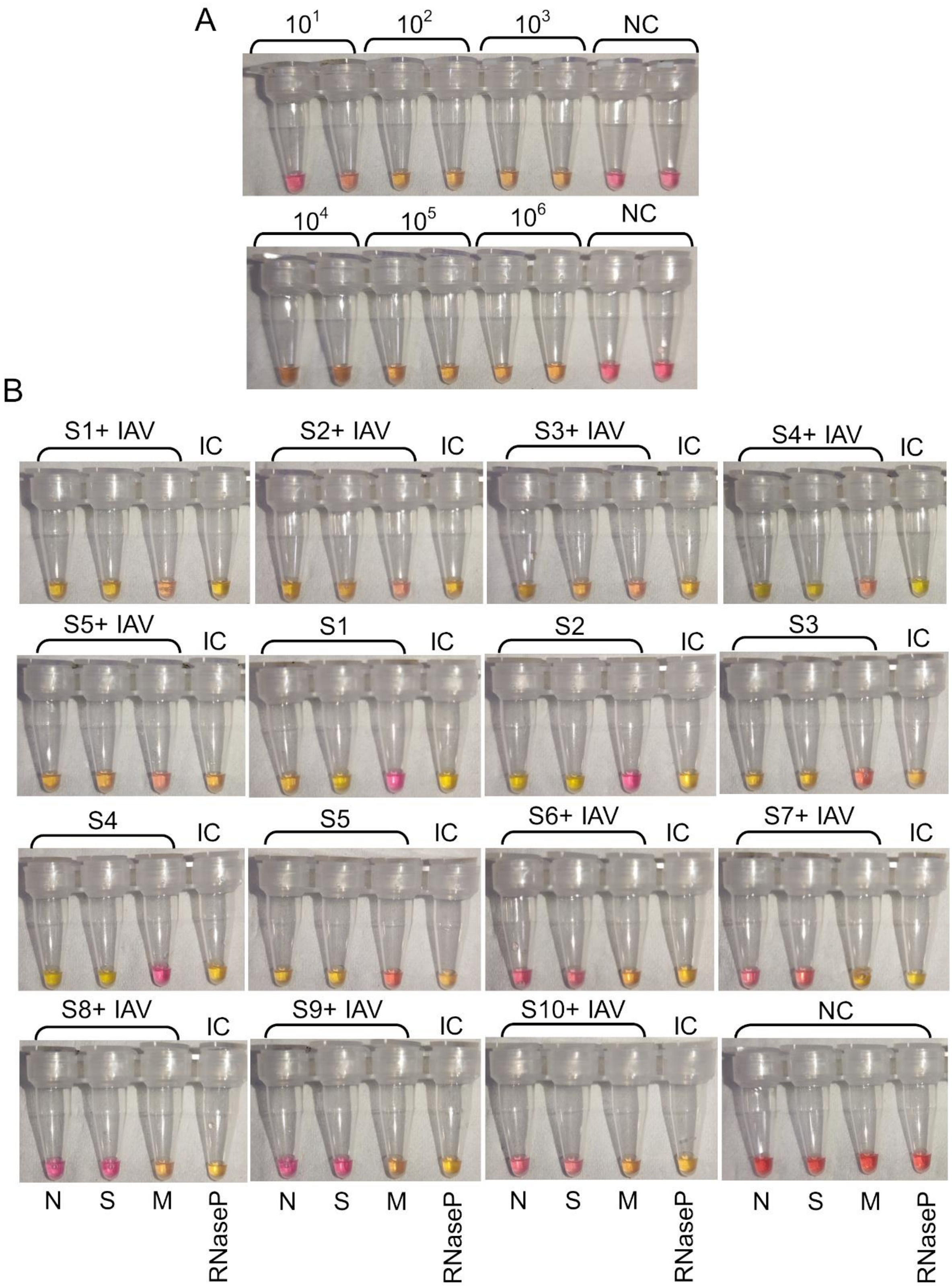


**Fig. S8:** (A) Colorimetric readout of limit of detection of the RT-LAMP method for detection Influenza A/H1N1/WSN/1933 virus. A known number of pfu/µL (dilutions containing $10^6$ to 10 pfu of virus per micro litre) of *virus* were amplified and detected by colorimetric RT-LAMP. (B) Colorimetric detection of co-infection using clinical SARS-CoV-2 samples having different Ct value spiked with 100 pfu of Influenza A/H1N1/WSN/1933 virus. IAV: Influenza A virus A, IC: Internal control, NC: Negative control
